# Technological advances in elite marathon performance

**DOI:** 10.1101/2020.12.26.20248861

**Authors:** Jonathon W. Senefeld, Michael H. Haischer, Andrew M. Jones, Chad C. Wiggins, Rachel Beilfuss, Michael J. Joyner, Sandra K. Hunter

**Author notes:** **Correspondence to:** Jonathon W. Senefeld, Ph.D., Department of Anesthesiology and Perioperative Medicine Mayo Clinic | 200 First Street SW | Rochester, MN 55905. JWS and MHH are co-first authors. MJJ and SKH have equal contributions and are co-senior authors.

## Abstract

There is scientific and legal controversy about recent technological advances in performance running shoes that reduce the energetic cost of running and may provide a distinct competitive advantage. To better understand the potential performance-enhancing effects of Nike’s pioneering marathon racing shoes, we examined the finishing times and racing shoes of the top 50 male and 50 female runners from the World Marathon Major series in the 2010s — before and after the introduction of new Nike shoe models (4%, NEXT%, Alphafly, and other prototypes; herein referred to as *neoteric Nikes*). Data for racing shoes were available for 3,886 of the 3,900 performances recorded at the four annual marathons in Boston, London, Chicago, and New York. In full cohort analyses, marathon finishing times were 2.0% or 2.8 min (138.5 ± 8.1 min vs. 141.3 ± 7.4 min, *P*<0.001) faster for male runners wearing *neoteric Nikes* compared to other shoes. For females, marathon finishing times were 2.6% or 4.3 min (159.1 ± 10.0 min vs. 163.4 ± 10.7 min, *P*<0.001) faster for runners wearing *neoteric Nikes*. In a subset of within-runner changes in marathon performances (males, *n* = 138; females, *n* = 101), marathon finishing times improved by 0.8% or 1.2 min for males wearing *neoteric Nikes* relative to the most recent marathon in which other shoes were worn, and this performance-enhancing effect was greater among females who demonstrated 1.6% or 3.7 min improvement (P=0.002). Our results demonstrate that marathon performances for world-class athletes are substantially faster wearing *neoteric Nikes* than other market-leading shoes, particularly among females.

## INTRODUCTION

The sub-2-hour marathon performance by Eliud Kipchoge (*1:59:40, hr:min:sec*) in late 2019 fascinated the public, athletes and scientists (7, 10, 12), not unlike the first sub-4-minute mile run by Sir Roger Bannister in 1954. This interest in the physiology of fast marathons is exemplified by the associated Viewpoint in *The Journal of Applied Physiology* (11) and ∼40 accompanying commentaries (22). Just hours after Kipchoge’s world best performance, the 16-year-old marathon world record for women was improved by 80 seconds by Brigid Kosgei. Kipchoge and Kosgei had one important commonality — both raced in a prototype in the latest line of marathon racing shoes from the Nike Vaporfly series. In a laboratory setting, the Nike Vaporfly 4% reduced the energetic of running among males by ∼4% relative to other contemporary racing shoes (5), hence the shoe’s moniker. These initial findings among males were supported and broadened to include females in independent laboratory testing (1) and analysis of real-world performance data of recreational runners (19). The Nike Vaporfly represented three deviations from “conventional” marathon performance shoes, each of which likely contributed synergistically to the 4% reduction in the energetic cost of running: first, embedded carbon-fiber plate (6, 21); second, innovative midsole material (13); and third, appreciable midsole thickness. This reduction in the energetic cost of running is predicted to improve running velocity to a lesser extent (∼2/3rds), thus, may improve marathon performance time by ∼2.5 minutes (14). However, translation of these laboratory findings to race performance among elite athletes has not been substantiated.

The unconventional Nike shoe models (4%, NEXT%, Alphafly, and other prototypes; herein referred to as *neoteric Nikes*) were originally developed in anticipation of the first widely-publicized attempt to break the 2-hour marathon barrier (Breaking2) held on May 6, 2017 (8). Akin to the whole-body polyurethane swimsuits used during the late 2000s to break over 100 world records, which were eventually banned from swimming competitions in 2010 (3), the *neoteric Nikes* led to widespread improvements in world records of distance road racing events and the introduction of new regulations for performance footwear in road running (2, 4). To better understand the performance-enhancing effects of the line of *neoteric Nikes* in real race settings, we examined elite marathon performances by athletes running with and without the *neoteric Nikes* in the World Marathon Major series.

The physiological requirements for fast marathon performances, including 2-hour marathon pace (10), are well-known and include an optimal combination of exceptional 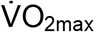, ‘lactate threshold’, and running economy (11). Although 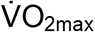 and ‘lactate threshold’ have been optimized by impressive training loads of elite athletes for many decades (11), improved running economy is thought to be deterministic in the most elite runners and is now the target of technological innovation to enhance human performance beyond current limits. Recent media articles suggested that the *neoteric Nikes* could improve marathon performance by ∼4% in sub-elite athletes (18). However, there is substantial heterogeneity between sub-elite marathon runners, and within-runner training may vary considerably particularly after the purchase of costly, exclusive *neoteric Nikes* with a perceived technological advantage. Thus, elite marathon runners are an ideal model to determine the effect of *neoteric Nikes* on marathon performance. Elite marathon runners are generally homogenous for consistent and intensive training across many years. Furthermore, among elite athletes, 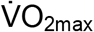 and running velocity at lactate threshold are generally stable across time and improvements in performance are primarily dependent on improved running economy which is typically improved gradually over many years, as demonstrated in a previous women’s world record holder for the marathon, Paula Radcliffe (9).

Accordingly, the objective of our study was to determine the relationship between marathon finishing times and the racing shoes worn by elite male and female marathon runners. This retrospective, observational study used real-world data (17) to test the *hypothesis* that marathon performances would be faster with *neoteric Nikes* for both cross-sectional and longitudinal observations. To accomplish this objective, we analyzed finishing times and the racing shoes worn by the top 50 males and females for four of the World Marathon Major races (Boston, Chicago, London, New York City) across a decade (2010 – 2019). Using the entire cohort, we compared marathon finishing times in elite runners with and without the *neoteric Nikes*. In a subset of elite runners with available repeat performances, including performances both with and without *neoteric Nikes*, we compared *within runner* changes in marathon performance. By focusing on elite performances, we also inherently controlled for any issues related to biological talent.

## MATERIALS AND METHODS

All procedures involved accessing public information and did not require ethical review as determined by the Mayo Clinic Institutional Review Board in accordance with the Code of Federal Regulations, 45 CFR 46.102, and the *Declaration of Helsinki*. Marathon finishing times and the associated racing shoes worn by the top 50 male and female finishers of the World Marathon Majors were collected.

The World Marathon Majors includes six of the largest and most renowned marathons in the world hosted by major cities (Berlin, Boston, Chicago, London, New York, and Tokyo). Tokyo and Berlin events were excluded due to the lack of marathon shoe data available for these competitions. Marathon finishing times were downloaded from Boston Athletic Association Archives (http://registration.baa.org/cfm_Archive/iframe_ArchiveSearch.cfm), Marathon Guide (London and TCS New York City Marathons; http://www.marathonguide.com/results/), and Bank of America Chicago Marathon Race Results (https://chicago-history.r.mikatiming.com/2018/).Finishing data from four races across 10 years of competition were collected and analyzed for the top 50 male and top 50 female finishers (except the New York City Marathon in 2012 which was not held due to the aftermath of Hurricane Sandy). Thus, a total of 3,900 data points were available for analysis.

Two of three investigators (JWS, MHH and RB) independently identified racing shoes as *neoteric Nikes* or other from available photographs posted on publicly available websites (e.g. https://www.marathonfoto.com/) or on social media webpages. Any disagreement was resolved by consensus of the three investigators. Of the 3,900 potential data points, 3,886 marathon performances (Male: 1,944; Female: 1,942) had identifiable shoes and were included in analyses.

For the subset analysis of elite runners with repeat performances, data were available for 1,505 performances (male, n = 799; female, n = 706). Of the 1,505 performances, 239 were completed in *neoteric Nike* shoes (male, n = 138; female, n = 101) and 1,266 were completed in other racing shoes (male, n = 661; female, n = 605).

Data were reported as means ± standard deviation within the text, unless noted otherwise. Changes in marathon finishing time between races for the case-control analysis were calculated as (race_n_ - race_n+1_) · (race_n_)^-1^ · 100%, where performances were arranged in chronological order and n = the first performance listed. Separate mixed-model univariate analyses of variance were used to compare the dependent variables (marathon finishing time and change in marathon finishing time) between the independent variables (sex, male vs. female; shoe, *neoteric Nike* vs. other; performance year; marathon course, Boston vs. Chicago vs. London vs. New York). Multiple comparisons tests were performed using the Bonferroni method (16). Analyses were performed with the use of IBM Statistical Package for Social Sciences version 25 statistical package (Armonk, NY, USA). Interpretation of findings was based on P < 0.05 or 95% confidence intervals. Reported P values are two-sided and have been adjusted for multiplicity using Bonferroni factor.

## RESULTS

### Full Cohort Analyses

Marathon finishing times were stable during the initial portion of the observation period (2010 – 2018) for both males (mean, 141.4 minutes; 95% confidence interval [CI], 141.0 to 141.7 minutes) and females (mean, 163.5; 95% CI, 162.9 to 164.0 minutes). In 2019, however, marathon performances markedly improved for both males (136.7 ± 0.4 minutes, P < 0.001) and females (158.3 ± 0.6 minutes, P < 0.001), **Figure 1**. Although this improvement in performance may be a characteristic of an Olympic qualification year, in the immediately preceding Olympic years (2015 and 2011), there was no change in performance for males (P = 1.0 for both) or females (P = 1.0 for both) relative to 2010, **Figure 1**. Each year, the average marathon finishing time was greater than 140 minutes for males and 160 minutes for females, except for 2019. The faster average marathon performance in 2019 coincided with the greatest proportion of males (n = 132 of 199 (1 missing observation); 66.3%) and females (n = 115 of 197 (3 missing observations); 58.4%) racing in *neoteric Nikes*.

**Figure 1.**
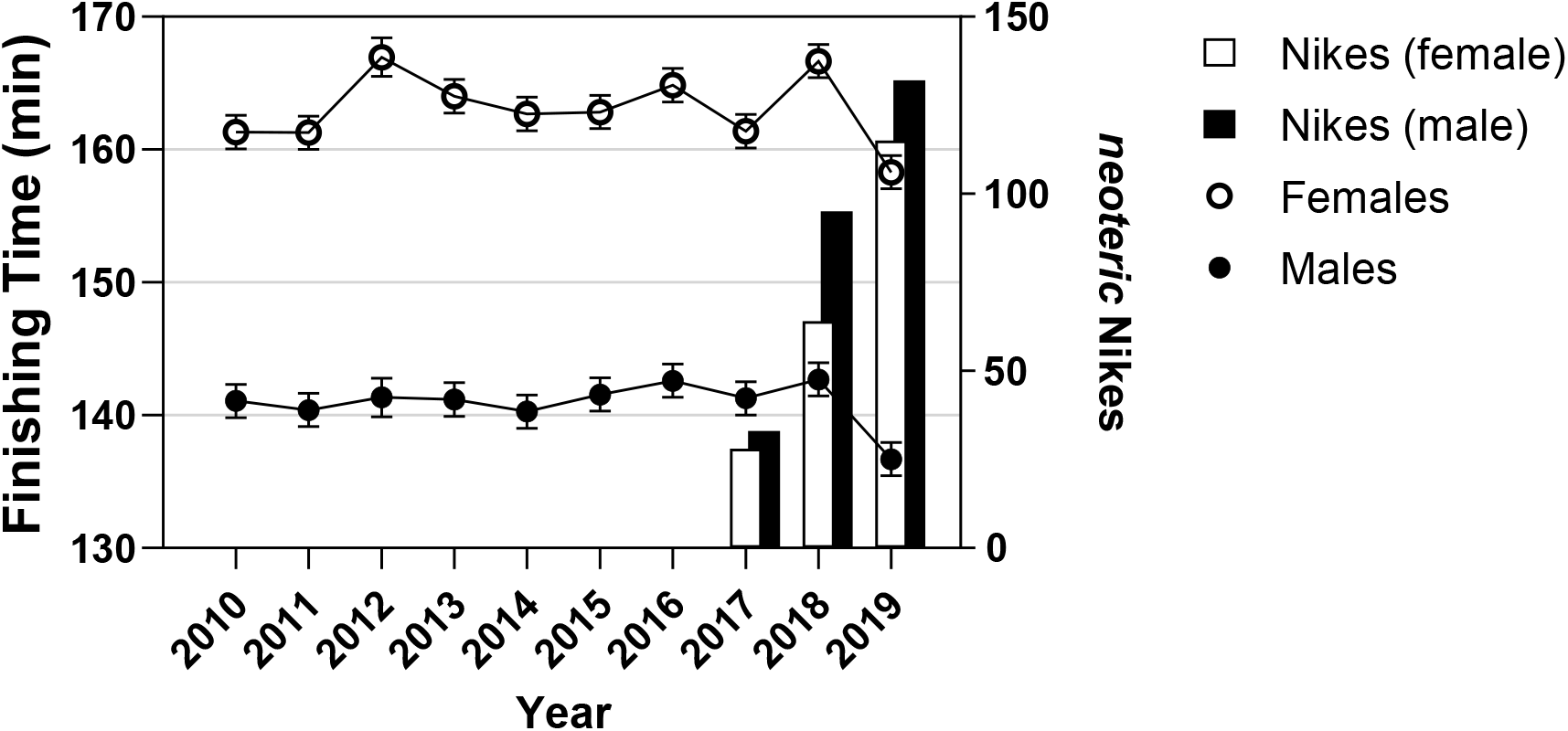
Finishing times of elite runners during the World Marathon Major races of the 2010s. The chronological dot plot represents the estimated marginal mean marathon performances times of males (**filled circles**) and females (**open circles**) who placed in the top 50 in a subset of World Marathon Major races (Boston, Chicago, London, or New York City) in the 2010s. The vertical error bars represent the 95% confidence intervals. For both males and females, the mean finishing time was faster in 2019 than all other performance years (P<0.001).

On average, the marathon finishing times of males were ∼22 min or ∼13% faster than the finishing times of females (141 ± 8 vs. 163 ± 11 min, P < 0.001). This finding was consistent with the marathon performances produced in other racing shoes (*n* = 3,419; P < 0.001). However, among the marathon performances in *neoteric Nike* shoes (n = 467), there were two primary differences.

First, marathon finishing times were faster among those runners wearing *neoteric Nike* shoes compared to other marathon racing shoes for both males (138.5 ± 8.1 min vs. 141.3 ± 7.4 min, P = 0.001) and females (159.1 ± 10.0 min vs. 163.4 ± 10.7 min, P<0.001), representing ∼2.0% faster performance among males and ∼2.6% faster performance among females. Second, the faster marathon performance in the runners wearing the *neoteric Nike* shoes compared to other marathon racing shoes was greater for females than males (P = 0.014; **Figure 2**). In the analytical model examining the modifying effect of sex and performance shoes on marathon finishing time, the effect size of the interaction of sex and shoe (P=0.014; η_p_ ^2^=0.002) was lower than the effect size of sex *per se* (P<0.001; η_p_ ^2^=0.341).

**Figure 2.**
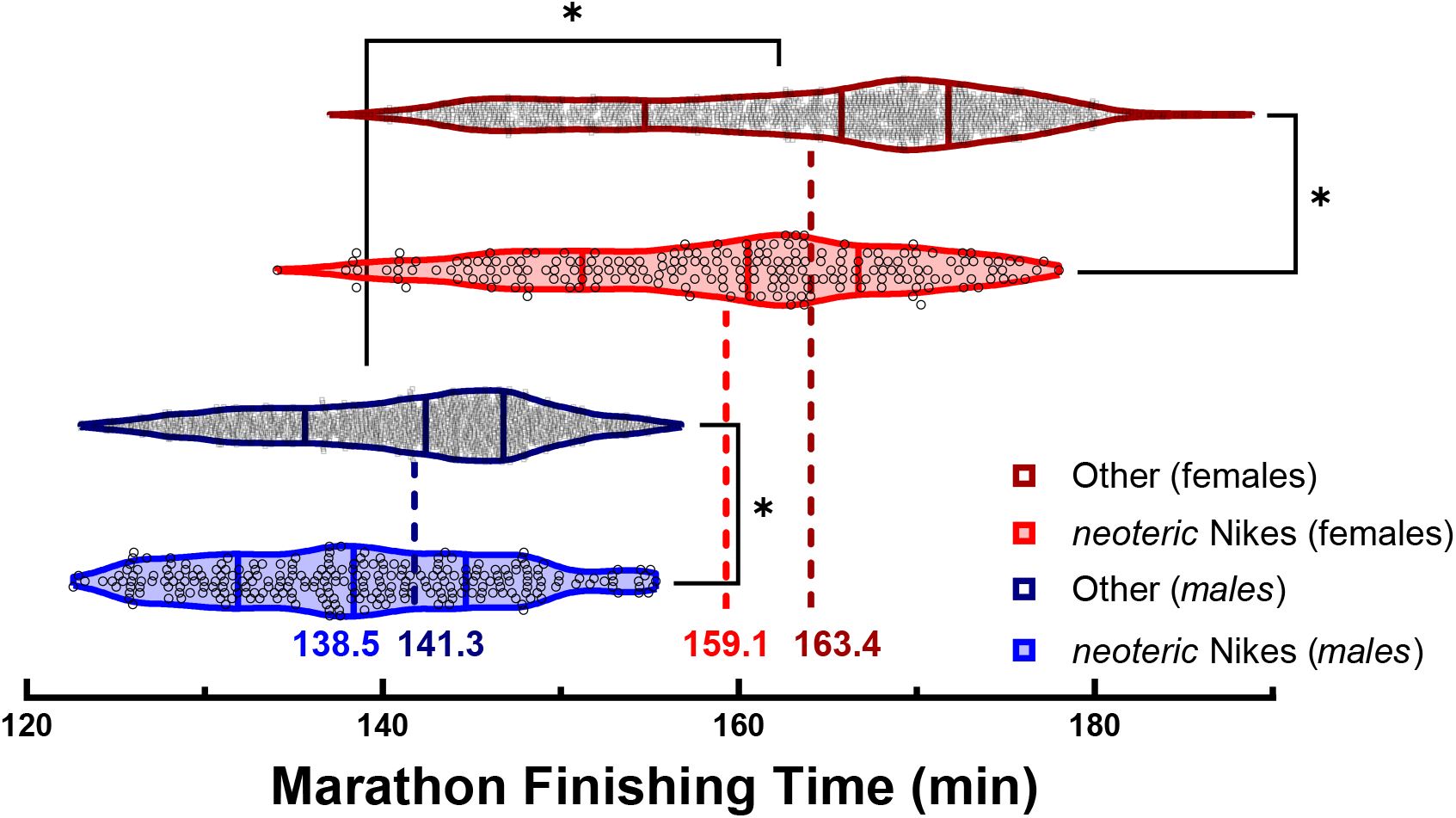
Finishing times in major marathons of elite athletes wearing *neoteric Nikes* or other running shoes. Violin plots represent the distributions of marathon finishing times of male and female athletes who placed in the top 50 in a subset of World Marathon Major races (Boston, Chicago, London, or New York City) in the 2010s. The middle vertical lines of each violin plot indicate the median, the left and right lines denote the 25^th^ and 75^th^ percentiles, and individual data points are indicated with open circles. Numerical values represent the mean finishing time of each distribution. For both males and females, marathon finishing times were faster among athletes wearing neoteric Nikes than athletes wearing other shoes; *, *P*<0.001.

### Case-Control Analyses of Repeated Performances

In a subset, case-control analysis of elite runners with repeat performances, we determined the within-runner change in finishing time between successive marathon performances wearing and not-wearing the *neoteric Nikes*. Consistent with our hypothesis, the change in performance between marathons was strongly moderated by racing shoe for both males (P<0.001) and females (P<0.001).

In the reference or control group of performances in non-Nike shoes, there was no observed between-race change in performance for males (median, - 0.0%; 95% confidence interval [CI], −11.7% to 12.4%) or females (median, 0.5%; 95% CI, −15.0% to 13.9%). Among these observations in the reference group, the probability of improvement in performance was 49.4% (327 events of 661 observations) for males and 59.2% for females (347 events of 605 observations). These analyses therefore, indicate no change in performance between successive marathon performances in the reference group (i.e. when wearing other racing shoes). See **Figure 3**.

**Figure 3.**
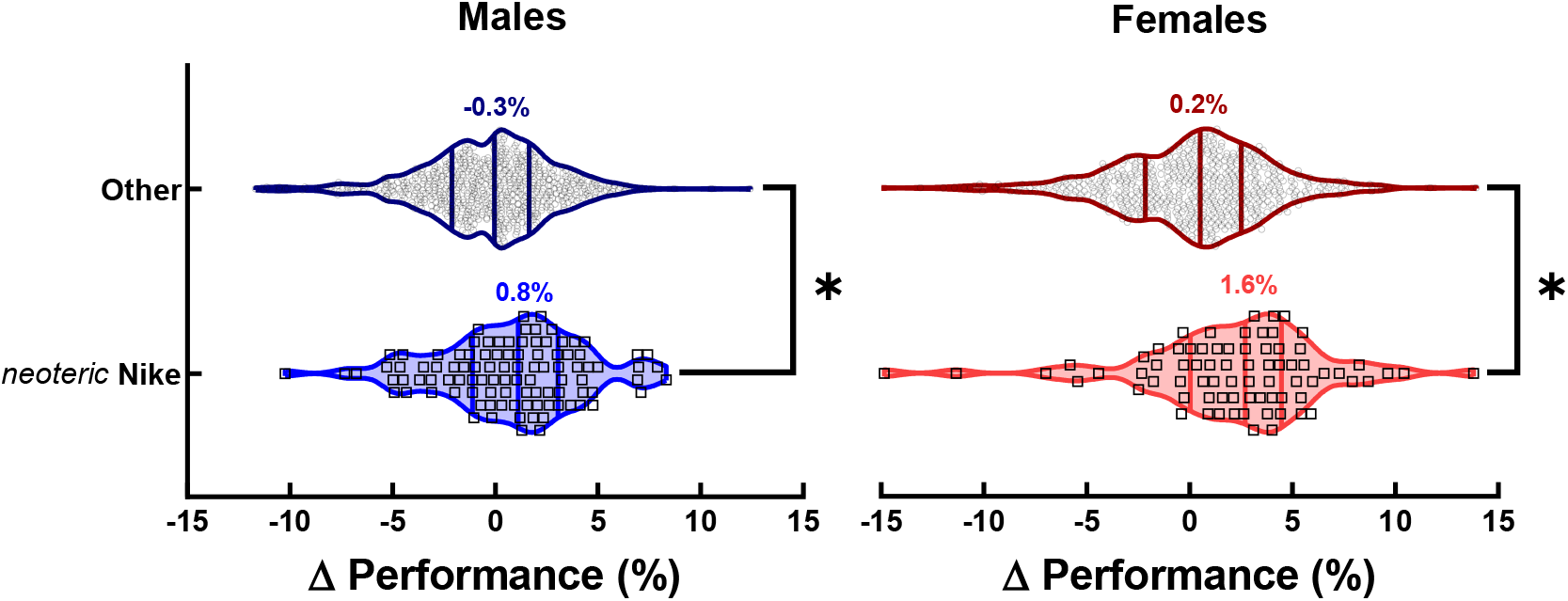
Within-runner successive marathon performances changes among elite athletes wearing *neoteric Nikes* or other running shoes. Distributions of race-to-race change in performance for males (left panel) and females (right panel) wearing non-*neoteric Nike* shoes (Other) or *neoteric Nike* shoes for the first time (after switch from other marathon racing shoe). Violin plots represent the distributions of marathon finishing times of male and female athletes who placed in the top 50 in a subset of World Marathon Major races (Boston, Chicago, London, or New York City) in the 2010s. The middle vertical lines of each violin plot indicate the median, the left and right lines denote the 25^th^ and 75^th^ percentiles, and individual data points are indicated with open circles. Numerical values represent the mean finishing time of each distribution. For both males and females, the change in marathon finishing times were improved among athletes wearing neoteric Nikes; *, *P*<0.001. However, marathon finishing times remain unchanged among athletes wearing other running shoes.

In the experimental or case group of performances in which *neoteric* Nike shoes were worn, the average between-race change in performance was 0.8% for males (median, 1.1%; 95% CI, −5.4% to 11.4%) and 1.6% for females (median, 1.8%; 95% CI, −6.9% to 13.8%). Among these observations in the experimental group, the probability of improvement in performance was 60.1% (84 events of 138 observations) for males and 70.3% for females (71 events of 101 observations). Thus, the relative risk of improvement in performance when wearing *neoteric* Nike shoes was 1.23 for both males and females. Although the relative change in performance (%) in *neoteric* Nikes was not different between the sexes (P = 0.158), the absolute change in finishing time (min) was greater for females than males (P < 0.001). These analyses demonstrate a performance-enhancing effect of *neoteric* Nikes, which was greater among females.

### Exploratory Analyses of Race course

Although the primary objective was to determine the association between marathon performance and the racing shoes worn, additional exploratory analyses were performed based on marathon race course. In support of other findings, marathon performance times of males were ∼10-13% faster than females for each marathon race course (all P < 0.001) and among marathon performances in other racing shoes (all P < 0.001). For each marathon race course, both males and females demonstrated faster performances wearing *neoteric Nikes* than other shoes (all *P*<0.001; **Figure 4**), with the exception of Boston for males (*P*=0.637). The null difference between shoe types for males in Boston likely reflects the difficult running conditions (heavy rain) during the 2018 Boston Marathon which slowed performances by six minutes on average. The 2018 Boston Marathon had high statistical leverage (*n* = 26; 45%) on the total sample of *neoteric Nikes* for males at Boston (*n* = 58). For females, however, the 2018 Boston marathon did not have large statistical leverage (*n* = 9; 27%) on the total sample of *neoteric Nikes* for females at Boston (*n* = 33), and thus did not notably affect these findings.

**Figure 4.**
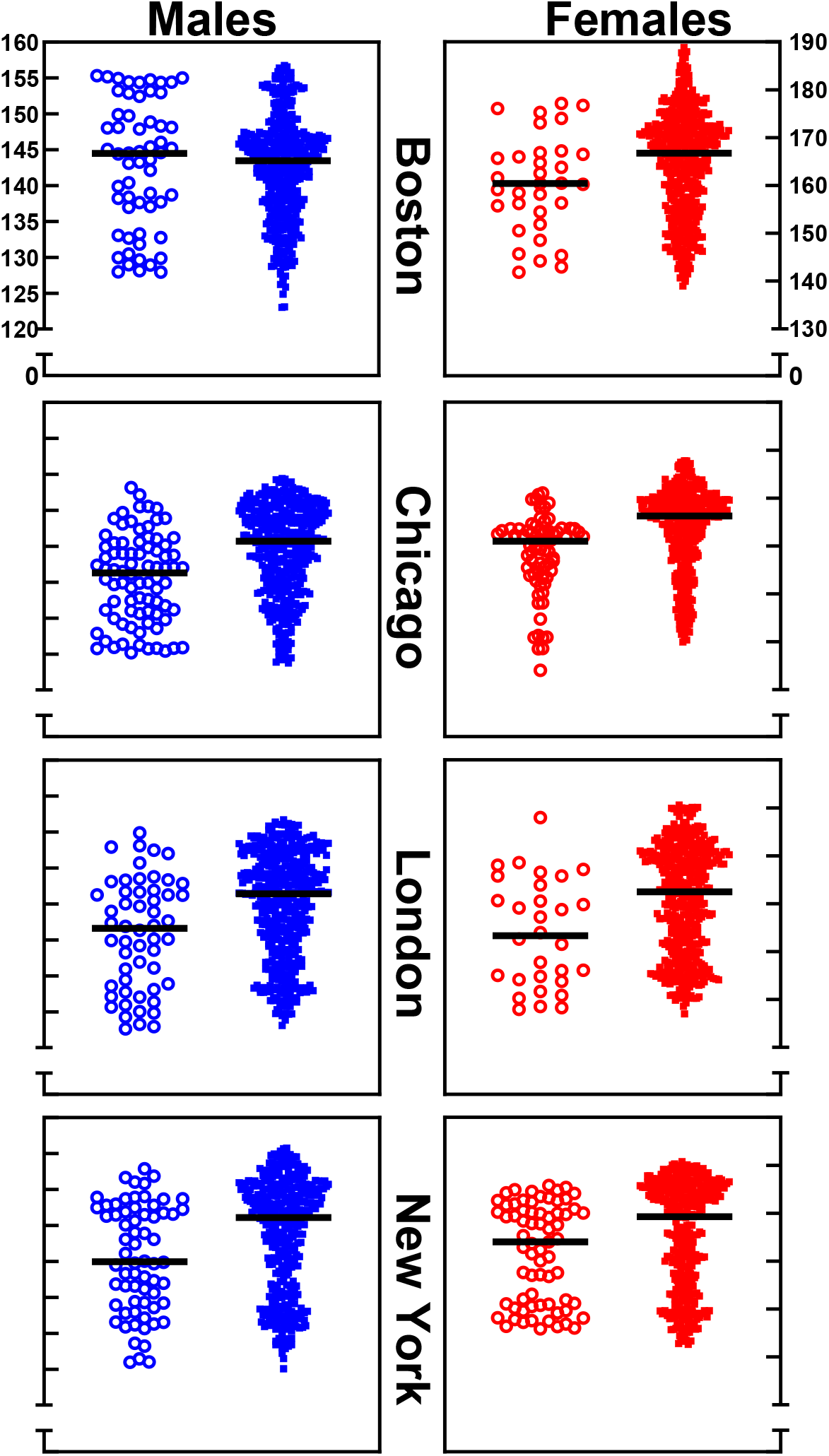
Race course specific marathon finishing times of elite athletes wearing *neoteric Nikes* or other running shoes. Distributions of marathon finishing times for males (left panels, blue symbols) and females (right panels, red symbols) for *neoteric Nike* shoes (unfilled symbols) and other shoes (filled symbols). The horizontal line in the middle of each distribution denotes the median of the sample.

## DISCUSSION

This retrospective, observational study using real-world data demonstrates a performance-enhancing effect of contemporary Nike marathon racing shoe models (4%, NEXT%, Alphafly, and other prototypes; *neoteric Nikes*) — which is greater for females than males. Our primary finding of performance-enhancing effects of *neoteric Nikes* compared other market-leading shoes, particularly among females, was supported by three separate analyses. First, the average marathon finishing time for both males and females was markedly faster in 2019 compared to previous years, which coincided with the first performance year in which the majority of runners wore *neoteric Nikes*. Second, marathon finishing times were faster among runners wearing *neoteric Nikes* compared to other shoes for both males (2.8 min or 2.0%) and females (4.3 min or 2.6%). Third, and perhaps most convincing, in a subset of elite runners with repeat performances, marathon finishing times improved for runners who switched to wearing *neoteric Nikes* relative to their most recent marathon wearing other shoes— for both males (0.8% or 1.2 min) and more so for females (1.6% or 3.7 min). Notably, there was no such change in marathon performance for males or females wearing other marathon racing shoes in repeated performances. These findings largely remained unchanged between different race courses in Boston, Chicago, London and New York. These findings suggest that technological advances in footwear contributed to the recent improvements in marathon performance times among elite runners and record-setting marathon performances.

The Nike Vaporfly 4% first became available to the public in late 2017. Since then, Nike has produced several iterations of the shoe with more refined characteristics of the innovative technology, i.e. embedded carbon fiber plate and thick midsole with novel foam material. Subsequently, this line of *neoteric Nikes* was worn by athletes to break world records in the marathon as well as other road races (100-km, half marathon, and 15-km distance) and has become almost omnipresent among eligible elite runners at marathons, including ∼70% of the top 50 males and females in the final World Marathon major race of the 2010s (New York Marathon held on November 3, 2019). The implementation of technology to improve the economy of movement has impacted nearly all modern-day sports (3, 4). Examples include introduction of carbon fiber and aerodynamic handlebars in cycling, clap skates in speed skating, ‘U’ grooves of club heads in golfing, fiber glass poles in pole vaulting, and ‘spaghetti strung’ rackets in tennis (3, 4). New technology introduced to sport elicits reconsideration and redevelopment of criteria to define the reasonable bounds of technological enhancement. In the case of long distance running, the *neoteric Nikes* motivated, in part, new regulations for performance footwear in road running (2, 4) after laboratory testing demonstrated a potential performance-enhancing effect (1, 5) and several world records were broken. Our findings support the notion that *neoteric Nikes* contributed to improvements in marathon finishing time in recent years for both males and females. Interestingly, the magnitude of improvements in performance are remarkably similar to the ∼2% faster performance predicted using models based on metabolic savings in running (14).

Although the relative (%) improvement in marathon performance was not different between the sexes, because males have faster performance times, the absolute improvement in performance wearing *neoteric* Nikes was numerically greater for females. This greater improvement in absolute running times of the females compared with the males was observed in the full-cohort analyses and also in the case-control data analyses of repeated marathon performances. Because marathon performances are determined in the time domain, as opposed to a relative performance, the greater benefit for females is noteworthy. Although the mass of the athlete and other biomechanical properties likely influence the performance benefit of the *neoteric Nike* shoes, there is limited empirical data evaluating the mechanisms contributing to potential sex-related differences in the performance benefit of the *neoteric Nike* shoes. Although most relevant performance prediction models do not directly account for the biological sex of the runner, analytical models predict that females would have a greater performance benefit due to slower performance times (14). The contributing mechanisms to the performance benefit of the *neoteric Nike* shoes, and particularly the potential sex-related differences, warrant further investigation.

Our findings are consistent with laboratory assessments of the *neoteric Nikes* (1, 5), analyses of real-world data from recreational marathon runners (19, 20), and analyses of elite athletes published in the lay literature (15). In the context of elite athletic performance, the observed ∼1.5% improvement in performance is substantial and highly meaningful for the elite-level athlete. For example, in the most recent Olympic Games in Rio de Janeiro in 2016, the top 10 men all finished the 42.2 km race within four minutes of one another. This improvement in performance is likely due to the elastic properties of the *neoteric Nikes* which conserves energy expenditure at marathon racing speeds (1, 5). Laboratory testing demonstrated that the *neoteric Nikes* could return 7.5 J of mechanical energy per step which is approximately double the energy return of other widely-used marathon performance shoes (3.5 J per step).

We conclude that the ingenious Nike performance running shoes with embedded carbon fiber plate and thick midsole with innovative material provide a distinct competitive advantage (∼1.5%) for both male and female elite marathon runners. Our findings indicate that the ∼4% reduced energetic cost of running observed in laboratory settings (1, 5) translates to real, but lesser, improvements in real world racing conditions among elite male and female marathon runners.

## Data Availability

All procedures involved accessing public information, as such, these data are publicly available.

## ACKNOWLEDGMENTS

None.

## AUTHOR CONTRIBUTIONS

JWS, MJJ, and SKH conceived and designed the study. JWS, MHH, CCW, and RB collected the data. JWS, MHH, and CCW analyzed the data and prepared the figures. All authors contributed to interpretation of results, drafting and revising the manuscript, and all authors approved the final version of the manuscript.

